## Supplementary material for "Sexually Transmitted Infections among Key Populations in India: A Protocol for Systematic Review": Search terms are provided in supplementary file no-01

**SUPPORTING FILE NO-01**

**Search terms** **and strategy**

A simple search strategy with search terms for PUBMED e-database are given as follows

**Search strategy**:

#1 AND #2 AND #3 AND #4

**Search terms**:

| **#4** | “India” |
| --- | --- |
| **#3** | (‘FSW’ OR ‘female sex workers’ OR SW OR sex workers OR prostitut* OR escort /OR commercial sex workers) OR (‘men who have sex with men’ OR men who have sex with men/ OR gay OR ‘homosexual male’/ OR homosexual* OR bisexual* OR ‘bisexual male’/ OR ‘men who have sex with men and women’/ OR ‘transsexual’ OR ‘transgender’ OR ‘TG’ OR ‘hijra’ OR kinnar OR kothis OR double-deckers OR panthis / OR ‘high risk population’ OR ‘high risk group population’/) OR (‘Transgender’ OR transgender male OR transgender female OR transgender population OR transgender people OR tg OR TG OR hijra/transgender OR H/TG OR HTG) OR (‘people with injecting drug use’ OR PWID OR IDU OR injecting drug user/s) |
| **#2** | (‘Sexually Transmitted Infection’ OR ‘sexually transmitted infection’ /OR ‘STI’ OR ‘Sexually Transmitted Disease’ OR ‘sexually transmitted disease’ /OR ‘STD’) (syphili* OR exp syphilis/ OR treponema pallidum OR Treponema pallidum/ OR great pox) OR (chlamydi* OR/ Chlamydia trachomatis OR chlamydia trachomatis OR C. trachomatis) OR (Neisseri* OR gonorrhoea*/ OR Neisseria gonorrhoeae OR neisseria gonorrhoeae) AND (Trichomona* OR Trichomoniasis/ OR Trichomonas vaginalis OR trichomonas vaginalis) |
| **#1** | Prevalence OR estimation |
