## Supplementary figures and images for "Sexually Transmitted Infections among Key Populations in India: A Protocol for Systematic Review"

### Data will be extracted by data extraction form

**SUPPORTING FILE NO-02**

**Data Extraction Form**

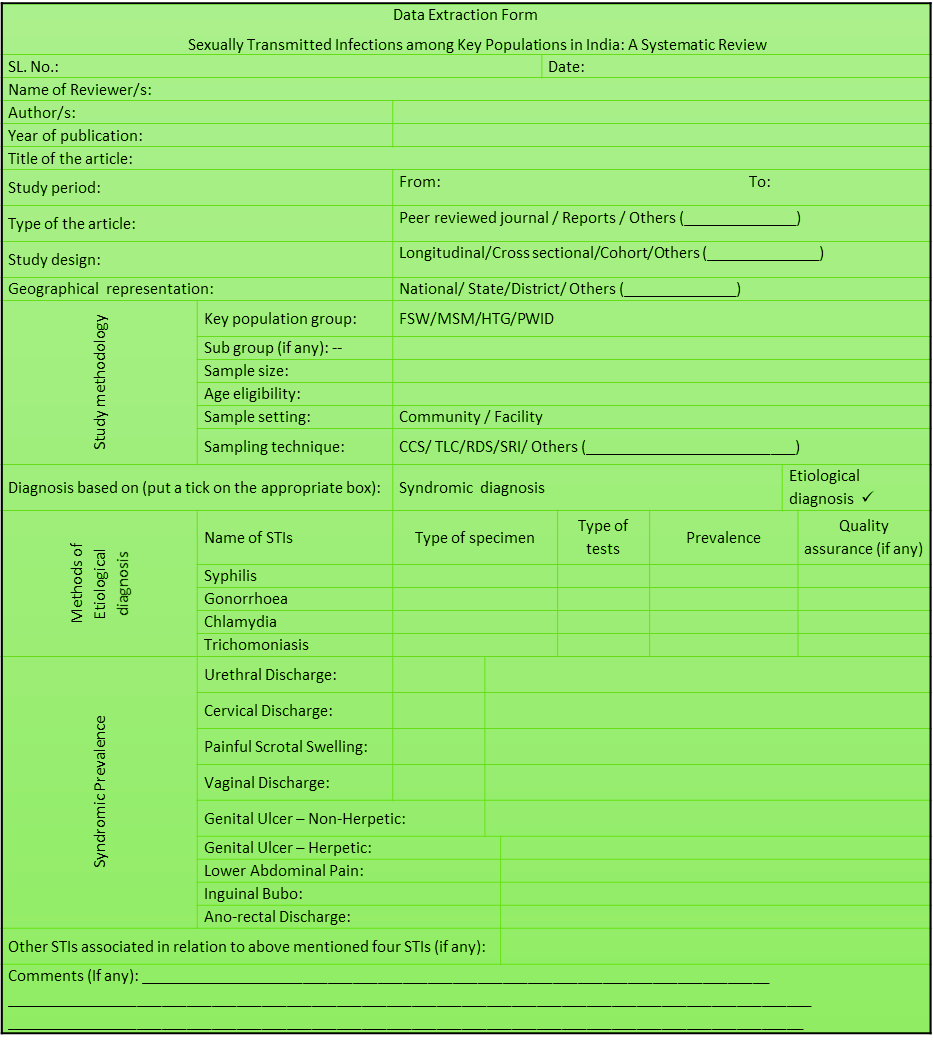
